# Self-Rated Health Inequalities Among Registered Nurses: A Cross-Sectional Analysis of UK Longitudinal Studies

**DOI:** 10.64898/2026.05.12.26352978

**Authors:** William P. Ball, Richard G. Kyle, Iain M. Atherton

## Abstract

**Background:** Health inequalities between occupational or social class groups are pervasive and persistent. Healthcare professionals have better health outcomes compared to the general population. Whether this is a result of healthcare education, favourable socio-demographic characteristics among professionals or other effects is not certain and the extent to which single healthcare occupational groups exhibit inequalities is unknown. We have described self-rated health and quantified geographic health inequalities among a single occupational group of Registered Nurses compared to the general population.

**Methods:** We analysed nationally representative samples from the 2011 UK Censuses across England, Wales and Scotland in the Office for National Statistics Longitudinal Study and Scottish Longitudinal Study. Self-rated health and socio-demographic characteristics for the study population are described. Inequalities in health by area deprivation among Registered Nurses and the General Population are quantified. Logistic regression analysis was used to assess the association between Nurse status and self-rated health, adjusting for socio-demographic variables.

**Results:** Among economically active, working age adults (n = 478,802), we identified 9,180 Registered Nurses resident in England, Wales and Scotland. 59% of Registered Nurses reported ‘very good’ self-rated health, with only 1% reporting ‘poor’ or ‘very poor’ health. A smaller proportion of Registered Nurses reported *less than good* health than the General Population at every level of area deprivation and had smaller absolute (4.1 percentage points vs. 9.1) and relative (RR: 1.5 vs. 2.0) inequalities between residents in the most and least deprived areas. Registered Nurses have an increased likelihood of reporting good or better health compared to the general population (Scotland – OR: 1.3, 95% CI: 1.2 – 1.5, England & Wales – OR: 1.4, 95% CI: 1.3 – 1.5) after adjusting for socio-demographic factors.

**Discussion:** Registered Nurses report better health compared to the general population and have smaller inequalities in health by area deprivation. However, unfair and avoidable geographical differences in health are present even in this socioeconomically privileged professional group. After adjusting for socioeconomic and demographic factors, the positive association between being a Registered Nurse and having good self-rated health remained.

## Introduction

Health inequalities, defined as systematic, unfair and avoidable differences in health outcomes between social groups, are persistent and widespread (McCartney et al., 2019). These inequalities have been observed across various measures of health and socioeconomic position (SEP), including occupation, education, and area-level deprivation. Despite an extensive body of research describing these inequalities, limited progress has been made in implementing effective policy to reduce them (Bambra et al., 2011; Kelly-Irving et al., 2022). The persistence of these inequalities, and in some contexts their continued growth (Bambra, 2024), highlights the importance of understanding the mechanisms through which SEP affects health so that we can design and evaluate effective policy interventions to address avoidable differences in health outcomes.

While there is a robust literature on the association between socioeconomic factors and health, much of this work relies on broad categorical measures such as occupational social class or area-based indices, which may fail to capture important within-group variation. For example, the assumption that all individuals within an occupational group, such as Registered Nurses (RNs), experience similar health outcomes may obscure inequalities shaped by geographic characteristics or individual-level circumstances. Conversely, reliance on aggregate geographic indicators may lead to the ecological fallacy, where assumptions are made about individuals based on area-level data. The complex interrelationships between individual and contextual SEP indicators remain insufficiently explored, particularly within occupationally defined populations.

This study addresses this gap by examining self-rated health (SRH) inequalities among Registered Nurses in Great Britain, using both occupational and geographic measures of SEP. By comparing RNs—a conventionally homogeneous occupational group—to the general population, this research explores whether health inequalities persist within occupational groups and how these relate to area-based deprivation. In doing so, it provides a foundation for understanding how SEP dimensions interact in shaping health outcomes and informs future policy development to reduce inequalities.

## Background

Research into health inequalities has consistently demonstrated that people from less advantaged socioeconomic groups experience worse health outcomes and shorter life expectancies than those from more advantaged backgrounds (Bambra et al., 2011; McCartney et al., 2019). The persistence of these inequalities across time, place, and population groups has made them a key focus of public health research and policy. Socioeconomic position is commonly operationalised using individual-level indicators such as education or occupation, and area-level indicators such as indices of multiple deprivation. Each approach has strengths and limitations, and many studies use them in combination to capture the multi-dimensional nature of SEP.

Research on health outcomes and behaviours among healthcare professionals presents a complex and varied picture. Studies have demonstrated that those with medical or nursing degrees have lower levels of self-reported morbidity and better overall wellbeing than individuals with other educational backgrounds (Shelton et al., 2021) and health-related behaviours among healthcare professionals are reportedly better than the general population (Schneider et al., 2018). However, occupational differences have been identified in health indicators such as obesity prevalence, which is higher among Nurses than the general population (Kyle et al., 2017, 2016), but lower for other healthcare professional groups. Explanations for these findings include the health-promoting effects of medical/healthcare education, occupational health benefits, and selection effects whereby healthier individuals are more likely to enter and remain in these professions. Moreover, healthcare professionals may possess greater health literacy and better access to services, which could further protect their health status. However, there is also concern around the healthcare work environment, increased experiences of stress and anti-social shift patterns (Holtzclaw et al., 2020).

Despite these general findings, there is a limited understanding of the extent to which health inequalities are present within single occupational groups such as Registered Nurses. Occupation-based classifications, such as the NS-SEC, often assume internal homogeneity, yet within-group variation—by income, geography, or working conditions—may produce meaningful differences in health. For example, all RNs are classified under the same Standard Occupational Classification (Office for National Statistics, 2016), but it remains unclear whether all members of this group experience uniformly better health or whether gradients persist within it.

Area-level deprivation measures are widely used in health research, but they are vulnerable to ecological fallacy. Not all individuals living in deprived areas are deprived themselves, and geographic measures may obscure the individual-level characteristics that contribute to poor health outcomes (Carstairs and Morris, 1989; Lokar et al., 2019). Conversely, focusing solely on individual indicators like occupation or education may overlook the broader structural and environmental factors that affect health. Although studies have begun to compare individual-level and aggregate measures (McCartney et al., 2023; Woods et al., 2022), few have examined how these intersect within specific occupational groups. There is a need for research that clarifies whether geographic inequalities in health persist even among those with presumed occupational advantages.

This study aims to address these gaps by conducting a stratified analysis of the health of Registered Nurses—a group typically considered socioeconomically advantaged—relative to the general population, and by examining the association between area deprivation and self-rated health within this group. Specifically, we use data from Census-derived data from across England, Wales, and Scotland to describe self-rated health among RNs. Logistic regression models are also employed to assess the association between RN status and SRH, adjusting for key socio-demographic characteristics.

Our research seeks to answer the following questions: 1) Do Registered Nurses exhibit health inequalities by geographic deprivation like those observed in the general population? 2) If so, are these inequalities of the same magnitude as observed in the general population? And 3) Is self-rated health associated with RN status after adjustment for socio-demographic characteristics? This research provides an important foundation for future research and policy work aimed at understanding and addressing health inequalities, even among groups who appear to be socioeconomically advantaged.

## Methods

### Design

This is a cross-sectional study utilising a representative sample of economically active working age adults from the 2011 Censuses for Scotland, England and Wales.

### Data

Individual level Census data was sourced from the Office for National Statistics Longitudinal Study (ONS LS) and the Scottish Longitudinal Study (SLS). The ONS LS contains linked administrative data from the National Census for England and Wales and other life events data, starting from 1971. It provides a largely representative 1% sample of the total population of England and Wales, with records for over 500,000 individuals at each Census time point included. Participants are included based on 4 randomly chosen birth dates, with updates to the sample at every Census time point. The Scottish Longitudinal Study (SLS) was developed to replicate the ONS LS. It is also based on the national Census for Scotland, but is a 5.3% sample of the Scottish population, with around 274,000 individuals initially selected from 1991 using 20 random birth dates.

ONS LS and SLS records were linked to two cross-national area deprivation measures using lower super output area (LSOA) and data zone small-area codes for place of residence. The adjusted UK Index of Multiple Deprivation (Abel et al., 2016) was constructed of scores from similarly measured domains of the separate Indices of Multiple Deprivation which cover the constituent countries of the UK which were normalised to a consistent scale. The Townsend 2011 deprivation measure (Yousaf and Bonsall, 2017) was constructed from scores based on consistently measured indicators of unemployment, car ownership, home ownership and household overcrowding in a given small-area across the United Kingdom.

### Study Population

The study population includes all individuals in the ONS LS and SLS samples from the 2011 Census time-point who were aged between 15 and 64 (inclusive) and were economically active (i.e. not long-term sick or retired based on the 2011 Census economic activity question). A sub-group of Registered Nurses were identified based on response to the Census occupation question and allocation to the Standard Occupational Classification (Office for National Statistics, 2016) group for Nurses (SOC code 2231). All other eligible individuals were classified as ‘general population’. Figure 1 outlines the inclusion/exclusion process and sub-groups.

**Figure 1.**
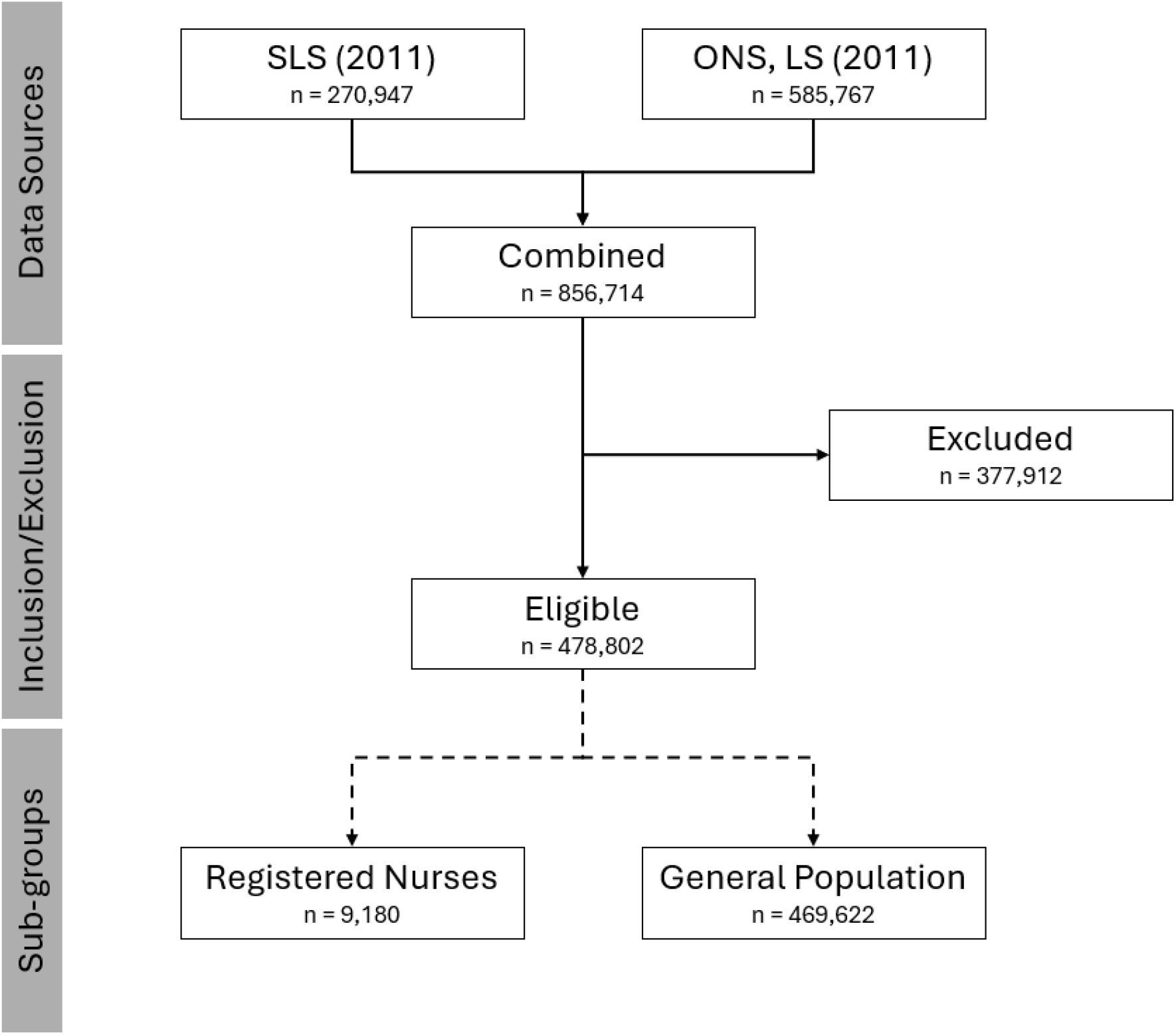
Study Population inclusion/exclusion flowchart

### Measures

Self-rated health (SRH) is a well-validated and widely used measure of overall health status, shown to be a strong independent predictor of mortality, morbidity, healthcare utilisation, and functional decline, even after adjustment for objective clinical indicators (DeSalvo et al., 2006; Idler and Benyamini, 1997). SRH captures aspects of physical, mental, and social wellbeing not fully reflected in clinical measures and has been used extensively in large-scale epidemiological studies and national surveys, including the UK Census (Jylha, 2009). Its inclusion in Census data makes it particularly suitable for population-level comparisons across occupational and geographic groups.

The main outcome of interest in this study is self-rated health measured from a Census question about general health. Respondents were asked ‘how is your health in general?’ with a five-point response scale - Very good, Good, Fair, Bad or Very Bad. A secondary outcome is sourced from responses to a question about health-related limitations. Respondents were asked ‘are your day-to-day activities limited because of a health problem or disability which has lasted, or is expected to last, at least 12 months?’, with a disclaimer prompting to ‘include problems related to old age’. The three responses available for this variable are ordinal; ‘Yes, limited a lot’, ‘Yes, limited a little’ and ‘No’.

Highest level of qualification was derived from a Census question which asks respondents to list all qualifications currently held and grouped into four categories for highest level of qualification and an additional level for no qualifications. Level 1 equates to school-leaving qualifications (e.g. GCSEs, O Levels or Standard Grades), Level 2 to pre-university qualifications (e.g. A Levels or Highers), Level 3 to post-school but pre-university qualifications (e.g. Higher National Certificate) and Level 4 to undergraduate or postgraduate degrees and professional qualifications (e.g. BA, MA, PhD, Medicine or Nursing).

### Statistical Analysis

#### Quantifying Inequalities

To meet our aim of assessing the presence and magnitude of socioeconomic inequalities in self-rated health among RNs and the General Population, we have calculated raw and population-weighted measures of absolute and relative inequality. We have calculated crude ranges of proportions reporting ‘less than good’ health between the most and least deprived areas, as well as Slope Index of Inequality (SII) and Relative Index of Inequality (RII) which can assess the full gradient (i.e. not only the most and least deprived) and account for different population sizes at each level of deprivation.

SII and RII quantify the absolute and relative differences in health between the most and least deprived areas. SII has been calculated separately for both RNs and the General Population using a linear regression method to account for the outcome value in each of the ranked deprivation quintiles, as well as population sizes (Low and Low, 2004). The outcome variable was the proportion of ‘less than good’ self-rated health responses at each deprivation quintile (measured by the dichotomous Self-Rated Health with Limitation measure described above). The independent variable was a relative rank variable calculated from the relevant population size and UK IMD quintile.

RII was calculated by dividing the SII for Registered Nurses and the General Population by their proportion reporting ‘less than good’ health for the overall population. RII can also be expressed as the percentage difference in an outcome between the most deprived area and the mean of the overall population (Low and Low, 2004).

Population Attributable Risk (PAR) is a measure which quantifies how much of a difference in an outcome between groups could be avoided in the hypothetical scenario that everyone belongs to the least disadvantaged group (Regidor, 2004). Using the least deprived UK IMD quintile as our reference group, we have calculated PAR to estimate how much of the difference in ‘less than good’ health could be explained by area deprivation.

#### The Association between Registered Nurse status and Self-Rated Health

A binary logistic regression model was calculated to quantify and assess the association between self-rated health and Registered Nurse status. The outcome variable, ‘Self rated health with limitation’, has been dichotomised from responses to the general health and health related limitation Census questions outlined above. ‘Good or better’ self-rated health (coded 1) is identified when the respondent answers Good or Very Good to the general health question and also answers No to the health-related limitation question. ‘Less than good’ self-rated health (coded 0) is identified when the respondent states that they have Fair or worse health in the general health question, or state that they have some limitation in response to the health-related limitation question. Registered Nurse status is derived from the Census occupation question and coded 0 (Non-Nurse) or 1 (Registered Nurse).

Demographic and socioeconomic covariates were selected based on the creation of a Directed Acyclic Graph outlining assumed relationships between the outcome, exposure and potential covariates. Covariates included in the model were age in years, sex and area deprivation for place of residence. We acknowledge the potential for additional unmeasured confounding and present this as a starting point for future causal analysis. Age was derived from the date of birth Census question and was calculated as the number of years between birth and Census day, 27^th^ March 2011. Sex was identified from response to a binary male/female Census question. Area deprivation was measured by the UK Index of Multiple Deprivation. The UK IMD ranks areas by a composite deprivation score (from most to least deprived) which are then grouped into deprivation quintiles.

We have reported odds ratios and 95% confidence intervals separately for England and Wales and for Scotland which represent estimates of the relative odds for Registered Nurses to report Good or Better self rated health compared to the general population in those longitudinal studies.

## Results

### Study Population Characteristics and Self-Rated Health

Among a total of 478,802 individuals matching our inclusion criteria in the combined ONS LS and SLS samples, we identified 9,180 Registered Nurses. Table 1 presents a summary of the study population and sub-group demographic, socioeconomic and health characteristics. Registered Nurses were older, with a mean age of 42.6 years (SD 10.7) compared to 39.2 years (SD 13) for the general population. Age distribution also differed between the two groups, with a smaller proportion of Registered Nurses aged below 40 years (38.6%) compared to the general population (50.3%). 88.7% of Registered Nurses were female, compared to 49.9% of the general population. A larger proportion of Registered Nurses (20.9%) were born outside of the United Kingdom compared to the general population (15.6%).

**Table 1.** Descriptive summary of study population for RNs and General Pop. Data Source: ONS LS and SLS.

| Variable | Level | Registered Nurses |  | General Population |  | Total |  |
| --- | --- | --- | --- | --- | --- | --- | --- |
|  |  | n | % | n | % | n | % |
| Age Groups | 15 – 24 | 425 | 4.6% | 82,176 | 17.5% | 82,601 | 17.3% |
|  | 25 – 29 | 790 | 8.6% | 51,854 | 11.0% | 52,644 | 11.0% |
|  | 30 – 34 | 1,050 | 11.4% | 50,204 | 10.7% | 51,254 | 10.7% |
|  | 35 – 39 | 1,281 | 14.0% | 52,164 | 11.1% | 53,445 | 11.2% |
|  | 40 – 44 | 1,423 | 15.5% | 56,870 | 12.1% | 58,293 | 12.2% |
|  | 45 – 49 | 1,597 | 17.4% | 58,206 | 12.4% | 59,803 | 12.5% |
|  | 50 – 54 | 1,365 | 14.9% | 50,723 | 10.8% | 52,088 | 10.9% |
|  | 55 – 59 | 848 | 9.2% | 41,092 | 8.8% | 41,940 | 8.8% |
|  | 60 – 64 | 401 | 4.4% | 26,333 | 5.6% | 26,734 | 5.6% |
| Sex | Male | 1,035 | 11.3% | 235,504 | 50.1% | 236,539 | 49.4% |
|  | Female | 8,145 | 88.7% | 234,118 | 49.9% | 242,263 | 50.6% |
| Country of Birth | Home LS | 6,464 | 70.4% | 360,358 | 76.7% | 366,822 | 76.6% |
|  | Other UK | 798 | 8.7% | 36,153 | 7.7% | 36,951 | 7.7% |
|  | Rest of World | 1,918 | 20.9% | 73,111 | 15.6% | 75,029 | 15.7% |
| Hours Worked per Week | ≤ 15 | 435 | 4.7% | 34,038 | 7.2% | 34,473 | 7.2% |
|  | 16-30 | 2,309 | 25.2% | 82,546 | 17.6% | 84,855 | 17.7% |
|  | 31-48 | 5,921 | 64.5% | 237,457 | 50.6% | 243,378 | 50.8% |
|  | ≥ 49 | 217 | 2.4% | 51,109 | 10.9% | 51,326 | 10.7% |
|  | NA | 298 | 3.2% | 64,472 | 13.7% | 64,770 | 13.5% |
| Housing Tenure | Owns | 7,178 | 78.2% | 310,011 | 66.0% | 317,189 | 66.2% |
|  | Other | 2,002 | 21.8% | 159,611 | 34.0% | 161,613 | 33.8% |
| Highest Level of Qualification | No Quals/<br>Missing/<br>Other | 185 | 2.0% | 89,935 | 19.2% | 90,120 | 18.8% |
|  | Level 1 | 165 | 1.8% | 88,134 | 18.8% | 88,299 | 18.4% |
|  | Level 2 | 242 | 2.6% | 82,021 | 17.5% | 82,263 | 17.2% |
|  | Level 3 | 317 | 3.5% | 68,248 | 14.5% | 68,565 | 14.3% |
|  | Level 4+ | 8,274 | 90.1% | 141,281 | 30.1% | 149,555 | 31.2% |
| UK IMD Quintiles | 5 – Least | 2,058 | 22.4% | 96,318 | 20.5% | 98,376 | 20.5% |
|  | 4 | 2,102 | 22.9% | 95,323 | 20.3% | 97,425 | 20.3% |
|  | 3 | 1,980 | 21.6% | 97,622 | 20.8% | 99,602 | 20.8% |
|  | 2 | 1,725 | 18.8% | 94,321 | 20.1% | 96,046 | 20.1% |
|  | 1 – Most | 1,315 | 14.3% | 86,038 | 18.3% | 87,353 | 18.2% |
| Townsend 2011 Quintiles | 5 – Least | 2,031 | 22.1% | 90,262 | 19.2% | 92,293 | 19.3% |
|  | 4 | 1,970 | 21.5% | 89,116 | 19.0% | 91,086 | 19.0% |
|  | 3 | 1,971 | 21.5% | 93,343 | 19.9% | 95,314 | 20.0% |
|  | 2 | 1,771 | 19.3% | 98,565 | 21.0% | 100,336 | 20.9% |
|  | 1 – Most | 1,437 | 15.6% | 98,336 | 20.9% | 99,773 | 20.8% |
| Self-Rated Health | Very Good | 5,402 | 58.8% | 243,117 | 51.8% | 248,519 | 51.9% |
|  | Good | 3,140 | 34.2% | 178,257 | 38.0% | 181,397 | 37.9% |
|  | Fair | 533 | 5.8% | 40,681 | 8.7% | 41,214 | 8.6% |
|  | Poor/Very Poor | 105 | 1.1% | 7,567 | 1.6% | 7,672 | 1.6% |
| Health-Related Limitation | A lot | 157 | 1.7% | 9,244 | 1.9% | 9,401 | 2.0% |
|  | Some | 487 | 5.3% | 27,976 | 6.0% | 28,463 | 5.9% |
|  | No Limitation | 8,536 | 93.0% | 432,402 | 92.1% | 440,938 | 92.1% |
| Self-Rated Health with Limitation | Less than Good | 930 | 10.1% | 62,462 | 13.3% | 63,392 | 13.2% |
|  | Good or Better | 8,250 | 89.9% | 407,160 | 86.7% | 415,410 | 86.8% |
|  | Total | 9,180 |  | 469,622 |  | 478,802 |  |

78.2% of Registered Nurses reported owning their home, either outright or with a mortgage/loan, compared to 66% of the general population. 9 in 10 (90.1%) Registered Nurses had their highest level of qualification in level 4 (degree and above, or professional registration), compared to less than a third (30.1%) of the general population. For both the UK IMD and Townsend 2011 measures, the general population had an almost even distribution of individuals living in each deprivation quintile. Registered Nurses were less likely to live in the most deprived areas (UK IMD – 14.3%, Townsend 2011 – 15.6%).

Registered Nurses reported higher proportions of ‘Very Good’ self-rated health (58.8% vs. 51.8%), and higher proportions of ‘Very Good’ or ‘Good’ self-rated health (93.0% vs. 89.8%) than the general population. Registered Nurses had a slightly smaller proportion who reported no health-related limitations (93.0% vs. 92.1%) compared to the general population, and a higher proportion with ‘Good or Better’ health in the dichotomised Self-Rated Health with Limitation variable (89.9% vs. 86.7%).

### Self-Rated Health Inequalities by Area Deprivation

Registered Nurses were less likely than the general population to report ‘less than good’ self-rated health with limitation at each UK IMD quintile (Figure 2 and Table 2). Social gradients in self-rated health with limitation were found for both Registered Nurses and the General Population, with larger proportions with poor health in the most deprived areas. Inequalities in the General Population were larger, with an absolute range of 9.1 percentage points in the proportion with ‘less than good’ health. The absolute range for Registered Nurses was smaller, with a 4.1 percentage point difference. The relative risk of reporting ‘less than good’ health in the most deprived areas compared to the least was 1.48 for Registered Nurses, and 1.96 for the General Population.

**Table 2.** Proportions of responses to dichotomised self-reported health for RNs and General Pop by area deprivation. Data Source: ONS LS and SLS.

|  |  | 5 - Least | 4 | 3 | 2 | 1 - Most |
| --- | --- | --- | --- | --- | --- | --- |
| Registered Nurses | Less than Good | 8.5% | 9.7% | 9.6% | 11.4% | 12.5% |
|  | Good or Better | 91.5% | 90.3% | 90.4% | 88.6% | 87.5% |
| General Population | Less than Good | 9.5% | 11.2% | 13.0% | 14.8% | 18.6% |
|  | Good or Better | 90.5% | 88.8% | 87.0% | 85.2% | 81.4% |

**Figure 2.**
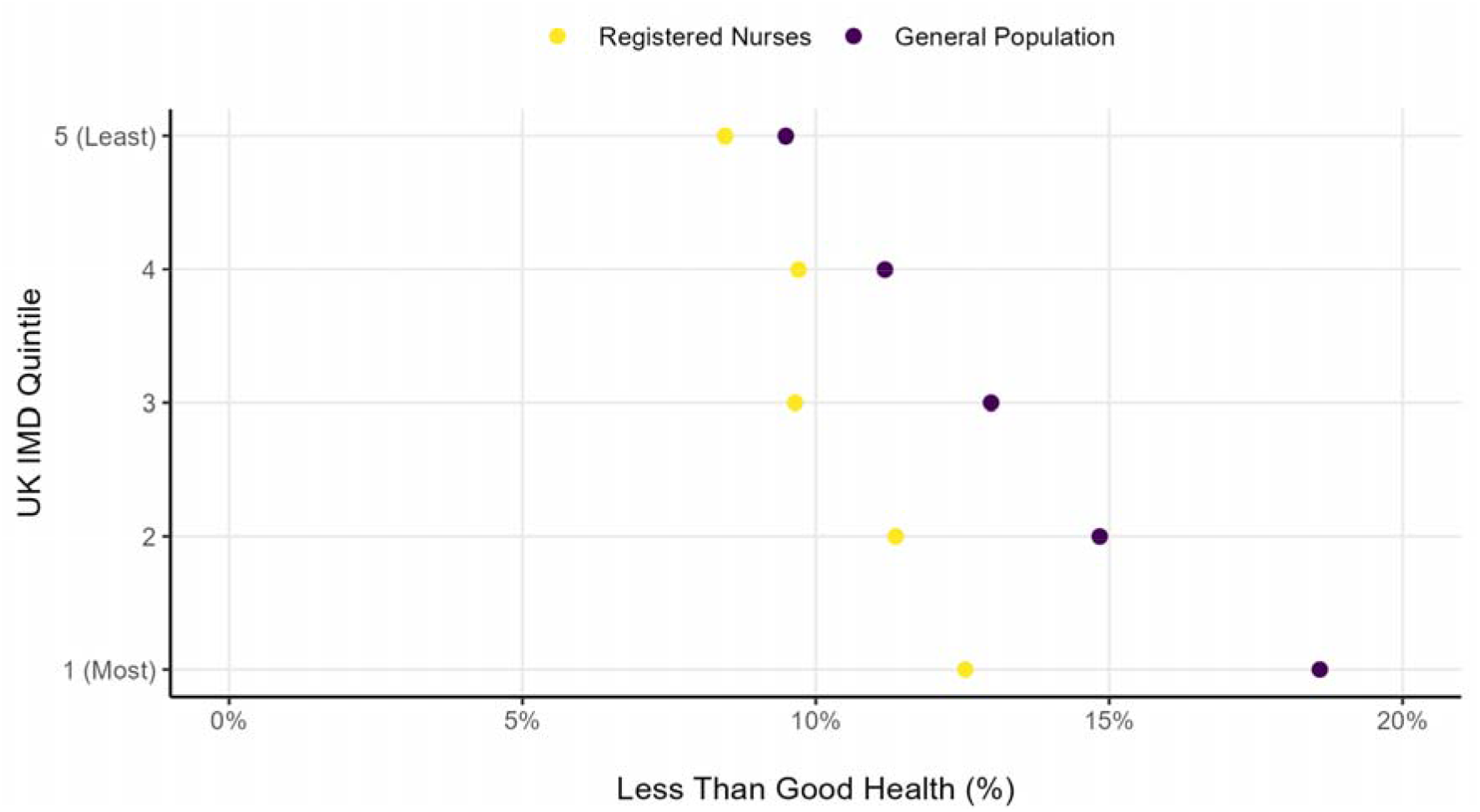
Proportions reporting less than good health by UK IMD for RNs and General Population. Source: Scottish Longitudinal Study and ONS Longitudinal Study

Slope Index of Inequality calculations also demonstrated that weighted absolute inequalities in the general population (SII: 0.043, i.e. +4.3 percentage points) are 3 times larger than for Registered Nurses (SII: 0.014, i.e. +1.4 percentage points). Relative Index of Inequality calculations estimate that for the general population, the most deprived group had a proportion reporting poor health that was 16.1% above the average for all levels of deprivation. For Registered Nurses, the most deprived group’s proportion was 6.8% above the average.

Area deprivation accounts for a larger proportion of inequalities in the General Population than in Registered Nurses. Based on Population Attributable Risk calculations, 16.5% of poor health responses among Registered Nurses were attributed to area deprivation, whilst this value was 28.7% for the General Population.

### Association between Nurse status and Self-Rated Health

In a logistic regression model adjusted for age, sex, and area-level deprivation (UK IMD quintile), registered nurses (RNs) had higher odds of reporting good or better health compared to the general population (Figure 3). The adjusted odds ratio (OR) was 1.33 (95% CI: 1.19–1.49) in Scotland, and 1.41 (95% CI: 1.32–1.52) in England and Wales.

**Figure 3.**
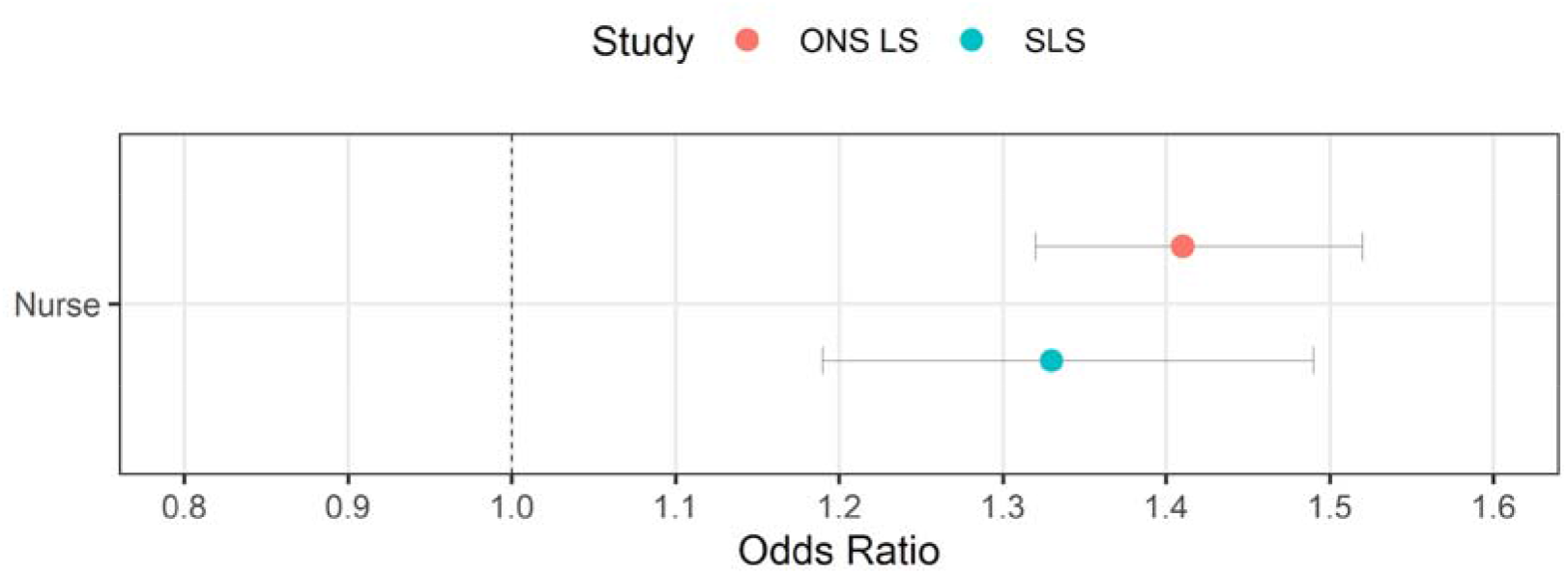
Odds Ratios for logistic regression models of the association between Nurse status and dichotomised self-rated health, adjusting for socio-demographic characteristics. Source: Scottish Longitudinal Study and ONS Longitudinal Study

## Discussion

### Principal Findings

We found that Registered Nurses have better self-rated health than the general population. On average, RNs are also older and more socioeconomically privileged. We also found that after adjusting for these sociodemographic differences, the association between positive self-rated health and RN status remained. We found that self-rated health inequalities by area deprivation are evident among RNs, but to a smaller extent than among the general population. Population Attributable Risk calculations revealed that area deprivation explains a smaller proportion of the inequality in self-rated health among RNs than the general population.

### Strengths and Limitations

This study benefits from the use of a large-scale representative sample from across Great Britain, drawn from the UK population Census which has a very high response rate. We also believe that this is the largest and most comprehensive study of its kind to explicitly explore health inequalities among RNs. We employ two distinct but generally concurrent area-based deprivation measures with different operationalisations/methodologies.

Our descriptive study is cross-sectional in nature and thus cannot (and does not attempt to) account for time-varying confounding or issues related to the temporal nature of causal relationships between our exposure and outcomes of interest. We do not estimate a causal effect size, but instead an association where residual confounding is likely and thus causal interpretations are not advised. In this analysis we rely on self-response to the Census occupation question to identify Registered Nurses and it is currently unknown how this compares to Nursing and Midwifery Council registration data, although known demographic characteristics of registrants largely match that of our self-identified sample (Nursing and Midwifery Council, 2017).

This study compares the self-rated health of Registered Nurses against a general population comparator which includes all other healthcare professional groups. Future work should examine other healthcare professional groups separately to enable more comprehensive comparative analyses across occupational cohorts. Such evidence would be valuable in informing targeted workforce interventions and policy priorities, including decisions about whether population health benefits may be maximised by prioritising specific professional roles

### How this study compares to other work

Self-rated health among Nurses and other healthcare professionals has been extensively studied, although rarely as the main outcome of interest and more likely as a predictor of other outcomes. Internationally, there is a high degree of variation in the proportion of Nurses reporting positive self-rated health, as demonstrated in Table 3 below.

**Table 3.** International studies measuring self-rated health among Nurses, by international context, level of positive health reporting and sample size.

| Study | International Context | Positive Health Proportion | Sample Size |
| --- | --- | --- | --- |
| (Cullati et al., 2017) | Switzerland | 93.5% | 466 |
| (Pien et al., 2019) | Taiwan | 82.8% | 1,690 |
| (Sveinsdottir and Gunnarsdottir, 2008) | Iceland | 78.5% | 394 |
| (Filha et al., 2013) | Brazil | 77.6% | 134 |
| (Gray et al., 2020) | Wales | 71.1% | 1,642 |
| (Pappas et al., 2005) | Greece | 41.6% | 353 |
| (Malinauskiene et al., 2011) | Lithuania | 39.6% | 748 |

Studies of nurses from Taiwan (Pien et al., 2019), Iceland (Sveinsdottir and Gunnarsdottir, 2008), Switzerland (Cullati et al., 2017) and Brazil (Filha et al., 2013) found very high levels of positive self-rated health, ranging from 70% to greater than 90% of respondents, which is in line with the findings of our study. Other work from around the world found very low levels of positive self-rated health, with those from Greece (Pappas et al., 2005) and Lithuania (Malinauskiene et al., 2011) finding the majority (58% and 60% respectively) of their samples reporting only ‘fair’ or worse health.

The previous Welsh study (Gray et al., 2020), based on 1,642 respondents - primarily nurses - found similar baseline characteristics to our study, including gender distribution (around 10% male), age distribution (fewer younger respondents and a peak above 40 years), and patterns of residential area deprivation. Nurses were more likely to live in less deprived areas, though nearly a quarter of participants had missing deprivation data, with unknown randomness. Most Welsh nurses reported good or very good health (71.1%) and no limiting illness (63.5%) – markedly lower than the 93.1% with positive self-rated health found in our project. Both the Welsh study and this project identified better health outcomes among residents in less deprived areas, though detailed cross-tabulations were not provided in the Welsh study.

Disparate international contexts are highly likely to vary due not only to differences in Nursing workforces’ sociodemographic factors, education and workplace conditions, but also how health varies between countries in general.

### Possible mechanisms and implications

Several mechanisms may explain the better self-rated health observed among Registered Nurses compared to the general population. Healthcare education may promote health literacy and health-protective behaviours, leading to earlier recognition and management of health problems. Selection effects may also play a role: healthier individuals may be more likely to enter and remain in nursing, while those with significant health limitations may divert to other occupations or leave the profession. Additionally, occupational benefits such as access to occupational health services, peer knowledge networks, and a generally stable employment environment may confer health advantages that persist even after adjusting for sociodemographic characteristics. The smaller geographic health inequalities observed among RNs compared to the general population may reflect the moderating effect of occupational advantage on area-level deprivation, whereby the professional and educational resources associated with nursing partially buffer the health-damaging effects of living in deprived areas. Nevertheless, the persistence of a socioeconomic gradient in self-rated health even within this privileged occupational group is important: it demonstrates that structural and environmental determinants of health operate independently of occupational status and cannot be fully offset by it.

These findings carry clear implications for workforce policy and health improvement strategy. Given that positive self-rated health is robustly associated with continued labour force participation, reduced sickness absence, and lower rates of early occupational exit, supporting nurses to maintain and improve their health is not merely a matter of individual wellbeing but a workforce sustainability imperative. The NHS 10 Year Health Plan for England (Department of Health and Social Care, 2025) explicitly commits to improving the working lives of NHS staff, including through a new set of Staff Standards that will outline minimum standards for modern employment and be reported at the employer level. Our findings suggest that attention to the geographic distribution of nursing staff, and the health conditions associated with residential deprivation, should feature in these standards and in workforce planning more broadly. Targeted occupational health support for nurses living in more deprived areas, where health risks are disproportionately concentrated, should be considered as they may help to retain experienced staff and reduce health-related attrition from the profession.

Our findings also point to the importance of widening access to nursing education and careers from more deprived areas. Given the strong association between RN status and positive self-rated health demonstrated here, expanding pathways into nursing for individuals from lower socioeconomic backgrounds could represent a meaningful lever for reducing health inequalities at the individual and household level. The NHS 10 Year Health Plan’s commitment to reorienting NHS recruitment towards its own communities, expanding apprenticeship routes, and supporting people from deprived backgrounds into training and entry-level roles provides a promising framework for this (Department of Health and Social Care, 2025). A nursing workforce that is more socioeconomically representative of the communities it serves may also be better positioned to understand and respond to the health needs of deprived populations, with potential downstream benefits for population health equity.

### Unanswered Questions and future research

This study is cross-sectional, meaning that it captures a single point in time and cannot address questions of causality or change. A longitudinal approach, following RNs and comparable non-nurse populations over time, would allow examination of how health trajectories diverge across careers, whether entry into nursing is associated with health improvements, and the extent to which occupational exit may bias cross-sectional comparisons. Longitudinal data would also enable analysis of whether geographic inequalities within the nursing workforce widen or narrow over time, and whether particular career stages or working conditions are associated with health decline.

More detailed socioeconomic information, particularly measures that capture early life circumstances, educational pathways, and household-level factors, would substantially strengthen future causal analyses. The current study is limited to the variables available in the Census, which, while broad, do not include income, parental occupation, childhood deprivation, or detailed working conditions. Future studies drawing on linked administrative data or cohort studies with richer socioeconomic measures could reduce residual confounding and facilitate the construction of more robust directed acyclic graphs. Additionally, examining whether the health benefits of RN status extend to household members or are concentrated at the individual level would help to clarify whether widening access to nursing careers could act as a route out of health disadvantage for families living in deprived areas. If future causal work were to confirm that nursing education and employment confer a direct health advantage, this would have important implications for widening access policy: expanding pathways into nursing for individuals from more deprived backgrounds, as envisaged in the NHS 10 Year Health Plan’s commitments to community-based recruitment and apprenticeship routes (Department of Health and Social Care, 2025), could represent a meaningful lever for reducing health inequalities at the individual level, in addition to its benefits for workforce sustainability.

## Declarations

### Ethical Approval

This study received ethical approval from the Edinburgh Napier University School of Nursing, Midwifery and Social Care Research Integrity Committee (Project Reference: FHLSS/1732).

### Data Availability

Underlying data used in this study can be accessed by application to the ONS Longitudinal Study and the Scottish Longitudinal Study, on condition that appropriate project approvals and accredited researcher status are secured.

### CRediT Authorship Contribution Statement

All authors (WB, IA, RK) were involved in the conceptualisation as well as review and editing of this article. WB wrote the original draft, conducted the formal analysis and investigation, curated data, wrote software, performed validation, produced visualisations, administered the project and acquired funding. Supervision was performed by IA and RK.

## Acknowledgements

The permission of the Office for National Statistics to use the Longitudinal Study is gratefully acknowledged, as is the help provided by staff of the Centre for Longitudinal Study Information & User Support (CeLSIUS). CeLSIUS is funded by the ESRC under project UKRI/ES/B001046/1. The authors alone are responsible for the interpretation of the data.

This work contains statistical data from ONS which is Crown Copyright. The use of the ONS statistical data in this work does not imply the endorsement of the ONS in relation to the interpretation or analysis of the statistical data. This work uses research datasets which may not exactly reproduce National Statistics aggregates.

The help provided by staff of the Longitudinal Studies Centre – Scotland (LSCS) is acknowledged. The LSCS is supported by the ESRC/UKRI the Scottish Funding Council, the Chief Scientist’s Office and the Scottish Government. The authors alone are responsible for the interpretation of the data. Census output is Crown copyright and supplied by National Records of Scotland. It is re-used under terms of the Open Government Licence.

## Appendices

### A. Model Output

Presented below are the full outputs for each logistic regression model used for analysis in this thesis. Model 1 was conducted separately for each Longitudinal Study (LS) and with an additional variable included as a sensitivity analysis. The eDatashield process does not provide a similar output and as the raw data is not shared with the analysis computer, there is no access to residuals. Further information is available (Raab et al., 2013).

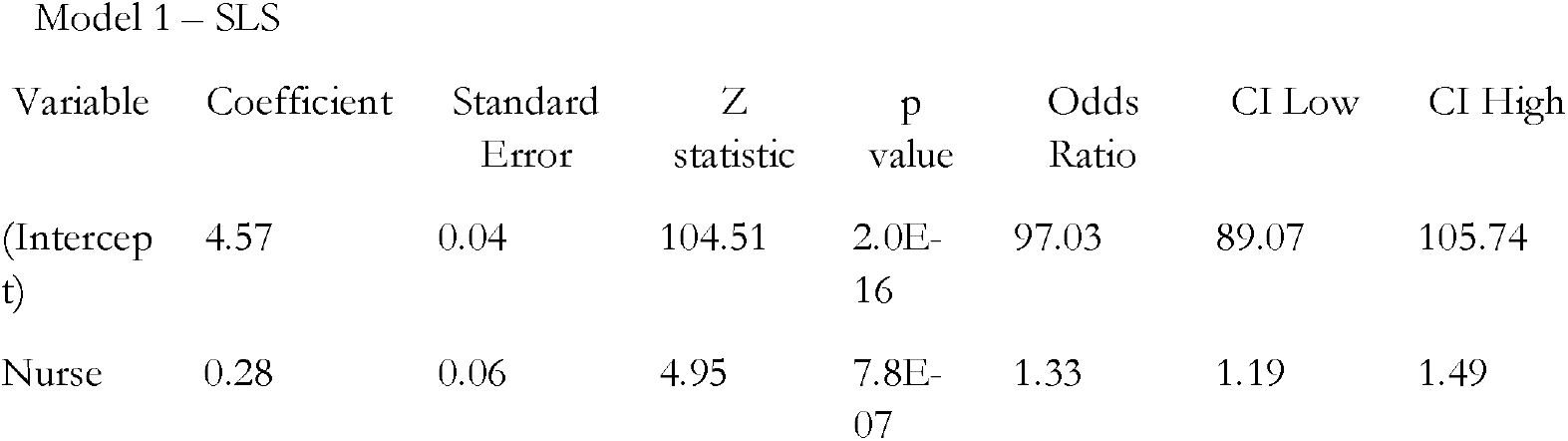
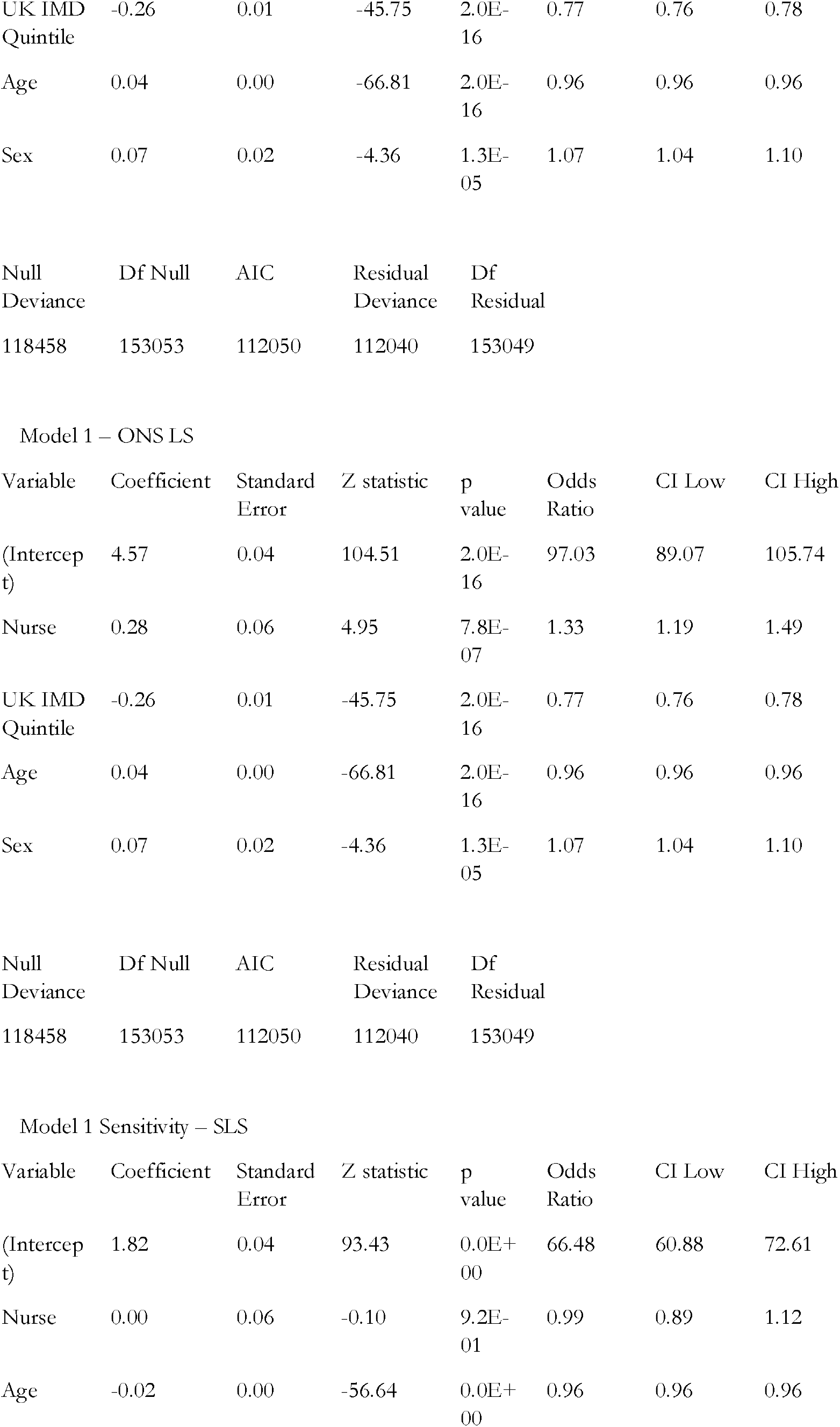
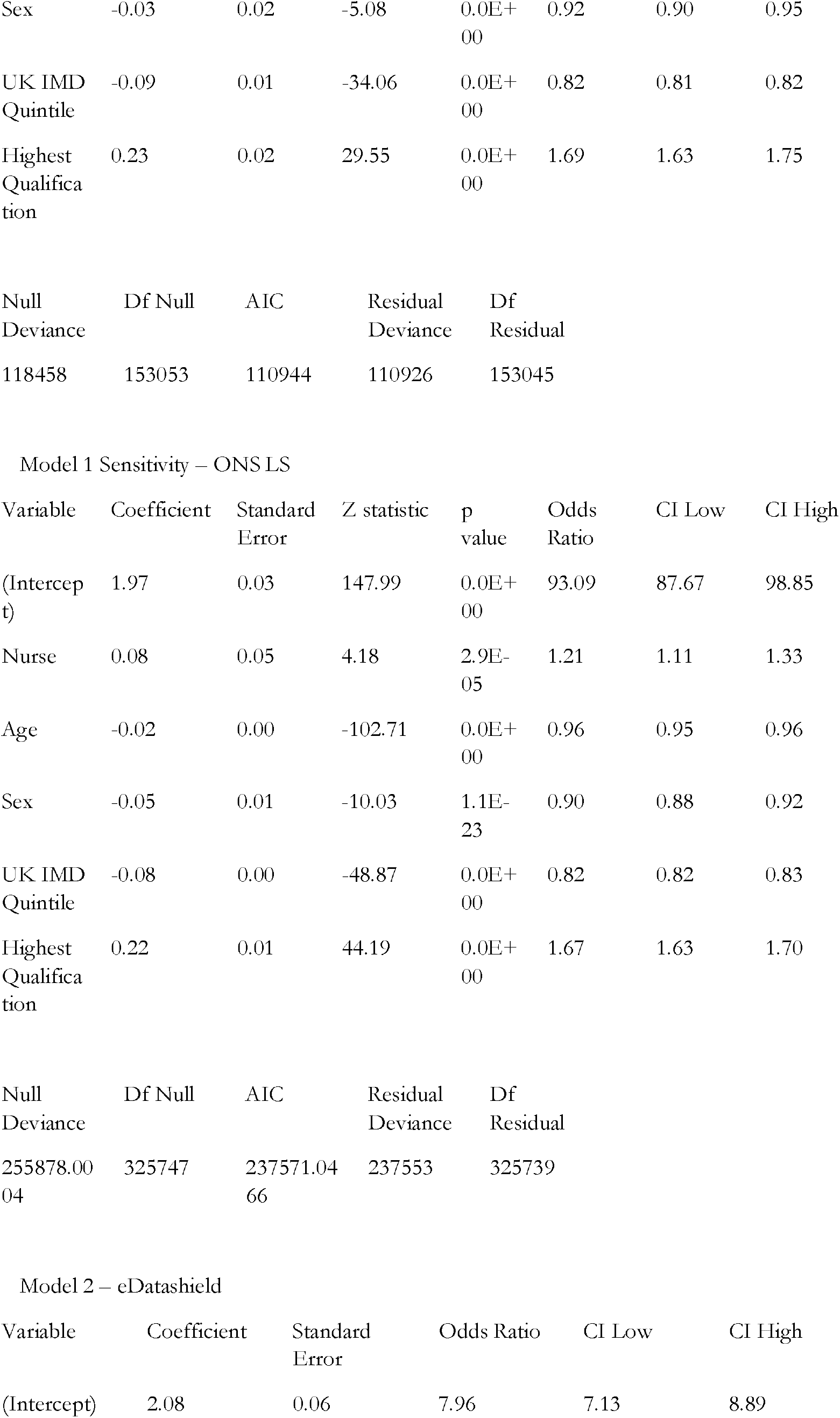
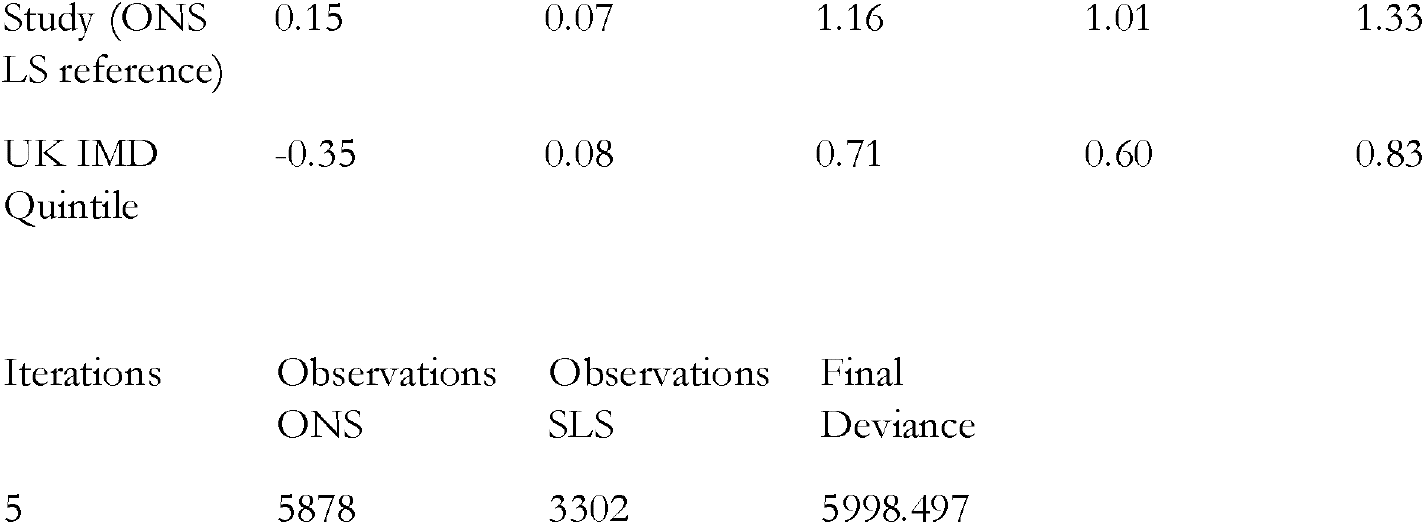

| Variable | Coefficient | Standard Error | Z statistic | p value | Odds Ratio | CI Low | CI High |
| --- | --- | --- | --- | --- | --- | --- | --- |
| (Intercept) | 4.57 | 0.04 | 104.51 | 2.0E-16 | 97.03 | 89.07 | 105.74 |
| Nurse | 0.28 | 0.06 | 4.95 | 7.8E-07 | 1.33 | 1.19 | 1.49 |
| UK IMD Quintile | -0.26 | 0.01 | -45.75 | 2.0E-16 | 0.77 | 0.76 | 0.78 |
| Age | 0.04 | 0.00 | -66.81 | 2.0E-16 | 0.96 | 0.96 | 0.96 |
| Sex | 0.07 | 0.02 | -4.36 | 1.3E-05 | 1.07 | 1.04 | 1.10 |

|  |  |  |  |  |
| --- | --- | --- | --- | --- |
| Null Deviance | Df Null | AIC | Residual Deviance | Df Residual |
| 118458 | 153053 | 112050 | 112040 | 153049 |

Model 1 – ONS LS
| Variable | Coefficient | Standard Error | Z statistic | p value | Odds Ratio | CI Low | CI High |
| --- | --- | --- | --- | --- | --- | --- | --- |
| (Intercept) | 4.57 | 0.04 | 104.51 | 2.0E-16 | 97.03 | 89.07 | 105.74 |
| Nurse | 0.28 | 0.06 | 4.95 | 7.8E-07 | 1.33 | 1.19 | 1.49 |
| UK IMD Quintile | -0.26 | 0.01 | -45.75 | 2.0E-16 | 0.77 | 0.76 | 0.78 |
| Age | 0.04 | 0.00 | -66.81 | 2.0E-16 | 0.96 | 0.96 | 0.96 |
| Sex | 0.07 | 0.02 | -4.36 | 1.3E-05 | 1.07 | 1.04 | 1.10 |

|  |  |  |  |  |
| --- | --- | --- | --- | --- |
| Null Deviance | Df Null | AIC | Residual Deviance | Df Residual |
| 118458 | 153053 | 112050 | 112040 | 153049 |

Model 1 Sensitivity – SLS
| Variable | Coefficient | Standard Error | Z statistic | p value | Odds Ratio | CI Low | CI High |
| --- | --- | --- | --- | --- | --- | --- | --- |
| (Intercept) | 1.82 | 0.04 | 93.43 | 0.0E+00 | 66.48 | 60.88 | 72.61 |
| Nurse | 0.00 | 0.06 | -0.10 | 9.2E-01 | 0.99 | 0.89 | 1.12 |
| Age | -0.02 | 0.00 | -56.64 | 0.0E+00 | 0.96 | 0.96 | 0.96 |
| Sex | -0.03 | 0.02 | -5.08 | 0.0E+00 | 0.92 | 0.90 | 0.95 |
| UK IMD Quintile | -0.09 | 0.01 | -34.06 | 0.0E+00 | 0.82 | 0.81 | 0.82 |
| Highest Qualification | 0.23 | 0.02 | 29.55 | 0.0E+00 | 1.69 | 1.63 | 1.75 |

|  |  |  |  |  |
| --- | --- | --- | --- | --- |
| Null Deviance | Df Null | AIC | Residual Deviance | Df Residual |
| 118458 | 153053 | 110944 | 110926 | 153045 |

Model 1 Sensitivity – ONS LS
| Variable | Coefficient | Standard Error | Z statistic | p value | Odds Ratio | CI Low | CI High |
| --- | --- | --- | --- | --- | --- | --- | --- |
| (Intercept) | 1.97 | 0.03 | 147.99 | 0.0E+00 | 93.09 | 87.67 | 98.85 |
| Nurse | 0.08 | 0.05 | 4.18 | 2.9E-05 | 1.21 | 1.11 | 1.33 |
| Age | -0.02 | 0.00 | -102.71 | 0.0E+00 | 0.96 | 0.95 | 0.96 |
| Sex | -0.05 | 0.01 | -10.03 | 1.1E-23 | 0.90 | 0.88 | 0.92 |
| UK IMD Quintile | -0.08 | 0.00 | -48.87 | 0.0E+00 | 0.82 | 0.82 | 0.83 |
| Highest Qualification | 0.22 | 0.01 | 44.19 | 0.0E+00 | 1.67 | 1.63 | 1.70 |

|  |  |  |  |  |
| --- | --- | --- | --- | --- |
| Null Deviance | Df Null | AIC | Residual Deviance | Df Residual |
| 255878.0004 | 325747 | 237571.0466 | 237553 | 325739 |

Model 2 – eDatashield
| Variable | Coefficient | Standard Error | Odds Ratio | CI Low | CI High |
| --- | --- | --- | --- | --- | --- |
| (Intercept) | 2.08 | 0.06 | 7.96 | 7.13 | 8.89 |

|  |  |  |  |  |  |
| --- | --- | --- | --- | --- | --- |
| Study (ONS<br>LS reference) | 0.15 | 0.07 | 1.16 | 1.01 | 1.33 |
| UK IMD<br>Quintile | -0.35 | 0.08 | 0.71 | 0.60 | 0.83 |

|  |  |  |  |
| --- | --- | --- | --- |
| Iterations | Observations<br>ONS | Observations<br>SLS | Final<br>Deviance |
| 5 | 5878 | 3302 | 5998.497 |

